# Hierarchical organ aging signatures from routine abdominal CT add incremental disease risk stratification beyond blood biomarkers

**DOI:** 10.64898/2026.05.19.26353206

**Authors:** Zengtian Deng, Yufeng Wang, Yu Shi, Lixia Wang, Touseef Ahmad Qureshi, Srinivas Gaddam, Sehrish Javed, Yin-Chen Hsu, Dante Rigo De Righi, Linda Azab, Garima Diwan, Ju Dong Yang, Yibin Xie, Chen Yuan, Camila Lopes Vendrami, Alex Rodriguez, Katherine Specht, Christie Y. Jeon, Humaira Chaudhry, James Buxbaum, Joseph R. Pisegna, Vahid Yaghmai, Wolfram Goessling, Yasmin G. Hernandez-Barco, Frank H. Miller, Temel Tirkes, Sara Espinoza, Nicolas Musi, Damini Dey, Kyung Hyun Sung, Stephen J. Pandol, Debiao Li

**Affiliations:** Biomedical Imaging Research Institute, Cedars-Sinai Medical Center, LA, CA, 90048, USA; Department of Bioengineering, University of California, Los Angeles, CA, 90048, USA; Department of Medicine, Samuel Oschin Comprehensive Cancer Institute, Cedars-Sinai Medical Center, Los Angeles, CA 90048, USA; Karsh Division of Gastroenterology and Hepatology, Cedars-Sinai Medical Center, 8900 Beverly Blvd, Los Angeles, CA 90048, USA; Comprehensive Transplant Center, Cedars-Sinai Medical Center, Los Angeles, CA 90048, USA; Samuel Oschin Comprehensive Cancer Institute, Cedars-Sinai Medical Center, Los Angeles, CA, USA; Department of Computational Biomedicine, Cedars-Sinai Medical Center, Los Angeles, CA, USA; Department of Radiology, Northwestern Memorial Hospital, Northwestern University Feinberg School of Medicine, IL 60611, USA; Division of Gastrointestinal and Liver Diseases, University of Southern California Keck School of Medicine, Los Angeles, CA 90033, USA; Division of Gastroenterology, Massachusetts General Hospital, Boston, MA 02114, USA; Biomedical Science Department, Cedars-Sinai Medical Center, Los Angeles, CA, 90048, USA; Department of Radiology, Rutgers New Jersey Medical School, NJ 07103, USA; Division of Gastroenterology, Hepatology and Parenteral Nutrition, VA Greater Los Angeles Healthcare System, CA 90073, USA; Radiological Sciences, University of California, Irvine CA 92868, USA; Department of Radiology and Imaging Sciences, Indiana University School of Medicine, IN 46202, USA; Center for Translational Geroscience, Department of Medicine, Cedars-Sinai Health Sciences Center, 8700 Beverly Boulevard, Los Angeles, CA 90048, USA; Division of Digestive and Liver Diseases, Cedars-Sinai Medical Center, Los Angeles, CA, 90048, USA

**Keywords:** biological age, abdominal CT, survival analysis, disease prediction, radiomics

## Abstract

Biological aging is heterogeneous across organ systems, yet whether CT-derived abdominal aging provides prognostic value beyond routine clinical data — and whether organ decomposition adds beyond a unified estimate — remains untested. We developed and evaluated organ-specific and ensemble biological age models from radiomic features across five abdominal organs in 68,682 CT scans from 32,882 subjects, evaluated on alignment with chronological age of healthy subjects (nested cross validation: MAE=3.68 years, R²=0.90). Age-interaction analyses showed attenuation of relative BAG–disease associations with advancing chronological age. We therefore performed focused prevention-oriented analyses in adults aged 20–60 years, the age range in which relative risk stratification was strongest. In this stratum, ensemble biological age gaps provided incremental prognostic value beyond demographic covariates for all-cause disease and mortality (ΔC-index=0.141, 0.051) and beyond routine blood biomarkers (ΔC=0.048), suggesting that CT-derived aging captures structural information complementary to selected blood markers. Organ-specific biological age added incremental prognostic value beyond ensemble selectively for focal diseases: cardiovascular (aorta, ΔC=0.091) and hepato-pancreatic (pancreas, ΔC=0.096). These findings establish a hierarchical organization of CT-derived biological aging, positioning routine CT as a source that adds prognostic value to existing clinical biomarkers.

## Introduction

Chronological age has long served as a practical reference for clinical risk assessment, but it is an incomplete measure of physiological aging. Individuals of the same chronological age can differ substantially in tissue integrity, organ function, and disease susceptibility. Seminal studies linking aging to cancer incidence [^1^] and chronic inflammation [^2^] further support the view that aging is a heterogeneous biological process rather than a uniform passage of time. This limitation has motivated the development of biological age biomarkers that quantify physiological aging beyond chronological time.

Biological age modeling has advanced across multiple biomedical domains. Epigenetic clocks estimate age from DNA methylation patterns and demonstrate that molecular aging can be measured across tissues [^3^]. Clinical and phenotypic clocks use routine laboratory and physiological measurements to capture morbidity and mortality risk beyond chronological age [^4,5^]. More recent work has extended biological age modeling to multi-organ molecular profiles, establishing that biological aging is a composite process across multiple organ systems and that organ-specific aging is disease-selective [^6–8^]. However, while these studies demonstrate that aging is heterogeneous across organs, a fundamental question remains unresolved: whether a unified aggregate biological age — collapsing multi-organ information into a single composite estimate — serves different clinical functions from organ-specific decomposition, and under what disease contexts organ-resolved aging provides prognostic value beyond aggregation. Furthermore, because most existing biological age models derive from blood, laboratory, or electronic health record (EHR) they cannot directly localize aging-related structural changes to specific tissues — a capability uniquely available through medical imaging.

Medical imaging offers a complementary route for biological age modeling by capturing anatomical and tissue-level manifestations of aging in vivo. MRI- and PET-derived brain age gaps have been associated with cognitive impairment, neurodegeneration, and Alzheimer’s disease, demonstrating that imaging-derived age deviations reflect clinically meaningful tissue degeneration [^9,10^]. Computed tomography is particularly attractive for systemic biological age modeling because it is performed opportunistically on tens of millions of patients annually in routine clinical care[^11,12^] — unlike dedicated MRI protocols or blood-based proteomics, CT-derived biological aging assessment requires no additional testing, no dedicated acquisition, and no added patient burden, positioning it as uniquely scalable for population-level aging biomarker deployment [^6^]. Prior CT studies have demonstrated feasibility of age prediction and associated CT-derived age gaps with disease and longevity [^13–15^]. However, whether CT-derived abdominal aging provides incremental prognostic value beyond available clinical data — including both demographic information and routine blood biomarkers — remains untested, leaving unresolved the unique clinical contribution of imaging-derived aging beyond what clinicians already measure.

Despite recent progress, four specific gaps limit the clinical translation of CT-based biological aging. First, existing CT aging models often lack large, strictly healthy, lifespan-spanning normative reference cohorts, limiting the ability to distinguish physiological aging patterns from occult or overt disease-related degeneration. Second, whether CT-derived abdominal biological aging provides incremental prognostic value beyond routine blood biomarkers has not been established, which is necessary to demonstrate that imaging-derived aging adds to the prognostic information already provided by systemic laboratory markers. Third, it remains unclear whether ensemble and organ-specific biological age gaps are clinically equivalent or hierarchically organized — specifically whether organ decomposition provides prognostic value beyond a unified abdominal estimate, and whether any such gain is general or selectively disease-dependent. Fourth, prior CT survival analyses have been conducted in general or screening populations without separating disease-mediated from mortality pathways less directly attributable to documented baseline disease, leaving unresolved whether CT-derived biological aging predicts mortality through mechanisms independent of diagnosed disease [^14,16^].

To address these gaps, we developed a radiomics-based abdominal CT biological age framework anchored to a large strictly healthy normative reference cohort spanning the adult lifespan. We trained organ-specific and ensemble abdominal age models and evaluated their hierarchical prognostic contributions through sequential incremental analyses — testing ensemble biological age against demographic covariates, organ-specific biological age against ensemble, and both against a baseline enriched with routine blood biomarkers — applied to pre-diagnostic scans obtained up to 10 years before first diagnosis and to mortality outcomes in the healthy reference cohort. We further examined biological plausibility by linking abdominal age gaps to routine blood-based biomarker measurements.

## Results

### Cohort construction and development of organ-specific abdominal biological age models

A total of 66,603 imaging scans from 30,818 subjects were collected from Cedars-Sinai Medical Center (in-house cohorts; Table S1). After filtering based on image quality and full existence of organs of interest without removal surgery, the whole data is split into the control cohort and the disease cohorts whose subjects ended up with at least one diagnosed disease based on Electronic Health Record (EHR). To further validate the robustness of the proposed age prediction model, two external cohorts were adopted in this study to further validate generalizability and translatability. One external clinical cohort consisted of 1959 contrast-enhanced CT scans and was used as a supplementary diagnostic stress test of PDAC-associated BAG elevation [^17^], while another cohort was multi-center control dataset with 120 scans collected from an R01 PDAC Prediction project. Detailed description of data cohort and demographic information can be found in Supplementary **Table S1.**

Within the 21,759 healthy scans shown in **Table S1**, 12,737 (58.5%) CT scans were utilized to train and validate the age prediction models, and the rest 9,022 (41.5%) scans served as the internal test dataset, all splits were done at subject level with stratification on chronological age, gender and body mass index (BMI). Based on **Figure 2d**, our proposed method achieved 3.68 years, 4.06 years, and 4.58 years in mean absolute error (MAE); 0.95, 0.92, and 0.88 in Pearson Correlation Coefficient (PCC); 0.90, 0.84, and 0.78 in R-Squared value (R2) for Nested Cross-Validation, internal test cohort, and external validation, respectively. As shown in **Figure 2c**, even on external validation control cohort, our proposed model’s predicted ages are well aligned with ground truth chronological age. We further benchmarked with the existing deep learning approach. We trained a deep learning network following the method of Kerber et al. on our in-house training cohort with 5-fold cross-validation [^13^]. Kerber’s method trained on our in-house dataset achieves better performance than their reported performance on our internal test dataset, with 4.90 years MAE, 0.89 PCC and 0.79 R^2^. However, our proposed method still outperforms Kerber’s method on our internal test dataset. The full benchmark comparison of more existing methods is shown in **Table S2**.

**Figure 1.**
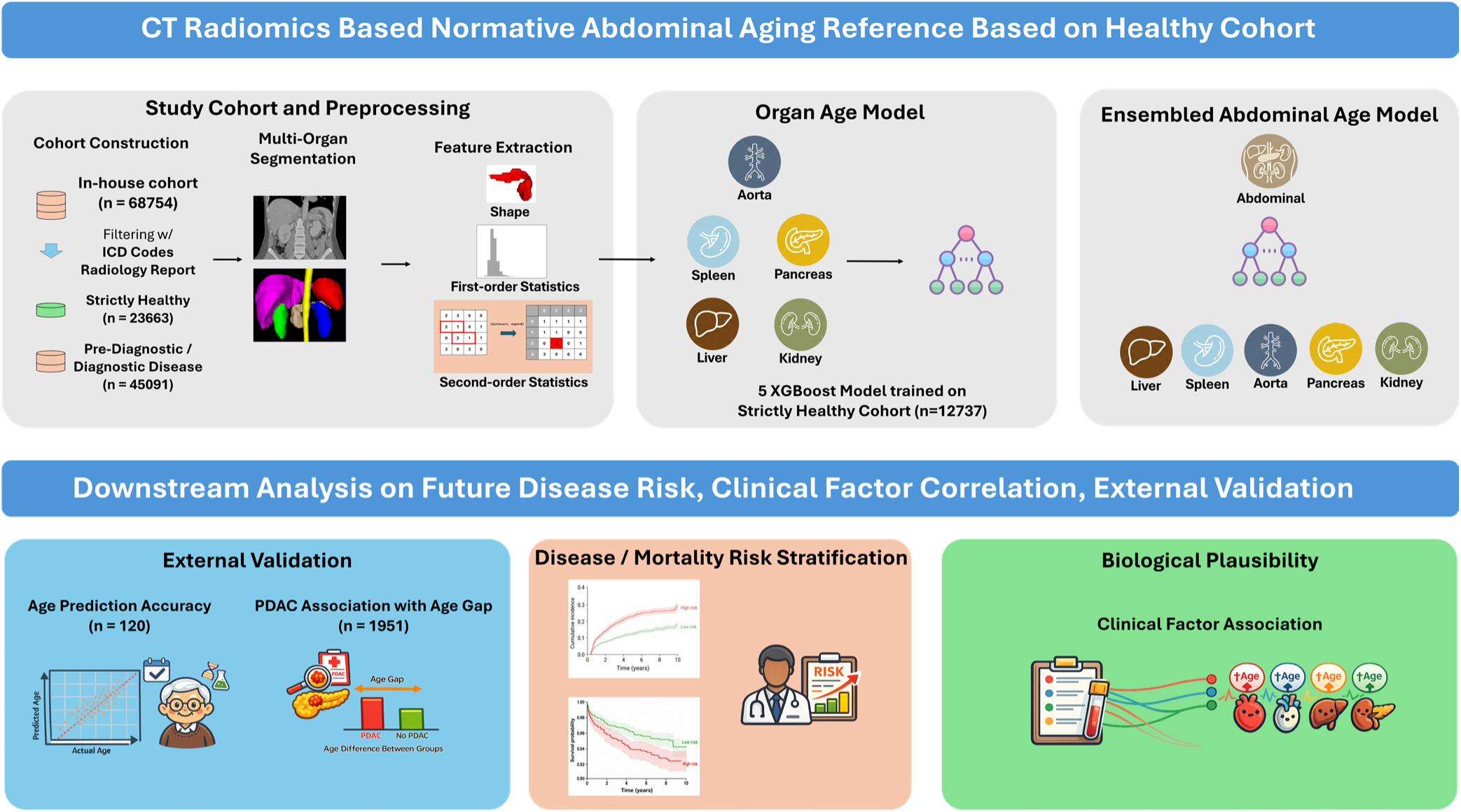
Overview of proposed Multi-Organ Biological Age Prediction Framework. In-house abdominal CT scans were filtered to define a strictly healthy cohort for model development and a separate pre-diagnostic/diagnostic cohort for downstream analyses. After multi-organ segmentation, shape, first-order and second-order radiomic features were extracted from the aorta, spleen, liver, pancreas and kidneys to train organ-specific XGBoost biological age models, which were further integrated into an ensemble abdominal biological age model. Organ-specific and ensemble biological age gaps were calculated relative to chronological age and used for downstream analyses of future disease and mortality risk stratification, routine blood biomarker associations, and external validation.

**Figure 2.**
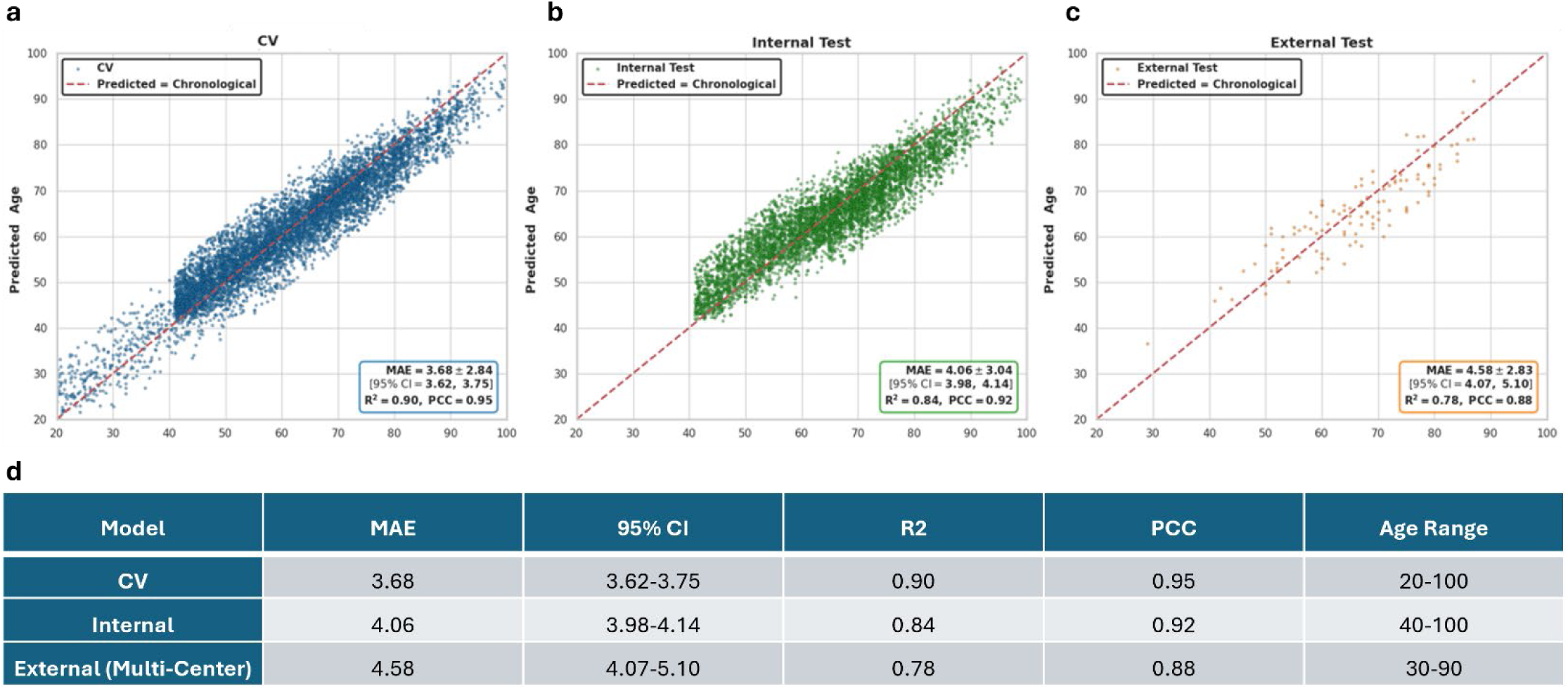
Age prediction performance of the ensemble abdominal biological age model across three evaluation settings. Scatter plots of predicted biological age versus chronological age for (a) nested 10-fold cross-validation (b) independent internal test cohort, and (c) external multi-center control cohort. Each plot shows individual scan predictions as colored dots; the dashed red line represents the identity line (predicted = chronological age). Performance metrics are annotated within each panel. (d) Summary table of MAE, 95% CI, R², PCC, and age range across the three evaluation settings.

### CT-derived abdominal biological aging is hierarchically organized for future disease risk stratification

We first evaluated whether chronological age modified the association between ensemble BAG and future disease risk. Age-interaction analyses showed attenuation of relative BAG-associated hazard with advancing chronological age, with the strongest relative associations in younger and midlife adults (Supplementary Figure S2). On this basis, the main risk-stratification analyses were focused on adults aged 20–60 years, a prevention-oriented subgroup in which relative BAG-associated risk was most pronounced. We performed Cox proportional hazards analysis using a mixed cohort of healthy and pre-diagnostic scans from subjects diagnosed with a disease of interest between 6 months and 10 years after imaging. Outcomes were grouped into four categories: cardiometabolic, hepato-pancreatic, GI cancer, and non-GI cancer. Biological age gaps (BAGs) were used as primary predictors following bias correction [6,9], with chronological age, sex, and BMI as covariates in all models.

Figure 3a shows that all organ BAGs except spleen yielded statistically significant HRs in the all-disease pooled analysis. At the disease-specific level, pancreas and aorta BAGs showed the highest HRs across most categories, while kidney BAG was elevated in cardiometabolic outcomes, liver BAG in heart disease and acute pancreatitis, and spleen BAG showed moderate associations with non-GI cancers. As shown in Figure 3b, all organ BAGs achieved significant ΔC over the demographic baseline, with the ensemble BAG performing best by integrating complementary organ-level signals. Per-disease analysis against the ensemble baseline (Figure 3c) further revealed clinically interpretable organ–disease associations, including aorta BAG for heart disease and spleen BAG for acute pancreatitis.

**Figure 3.**
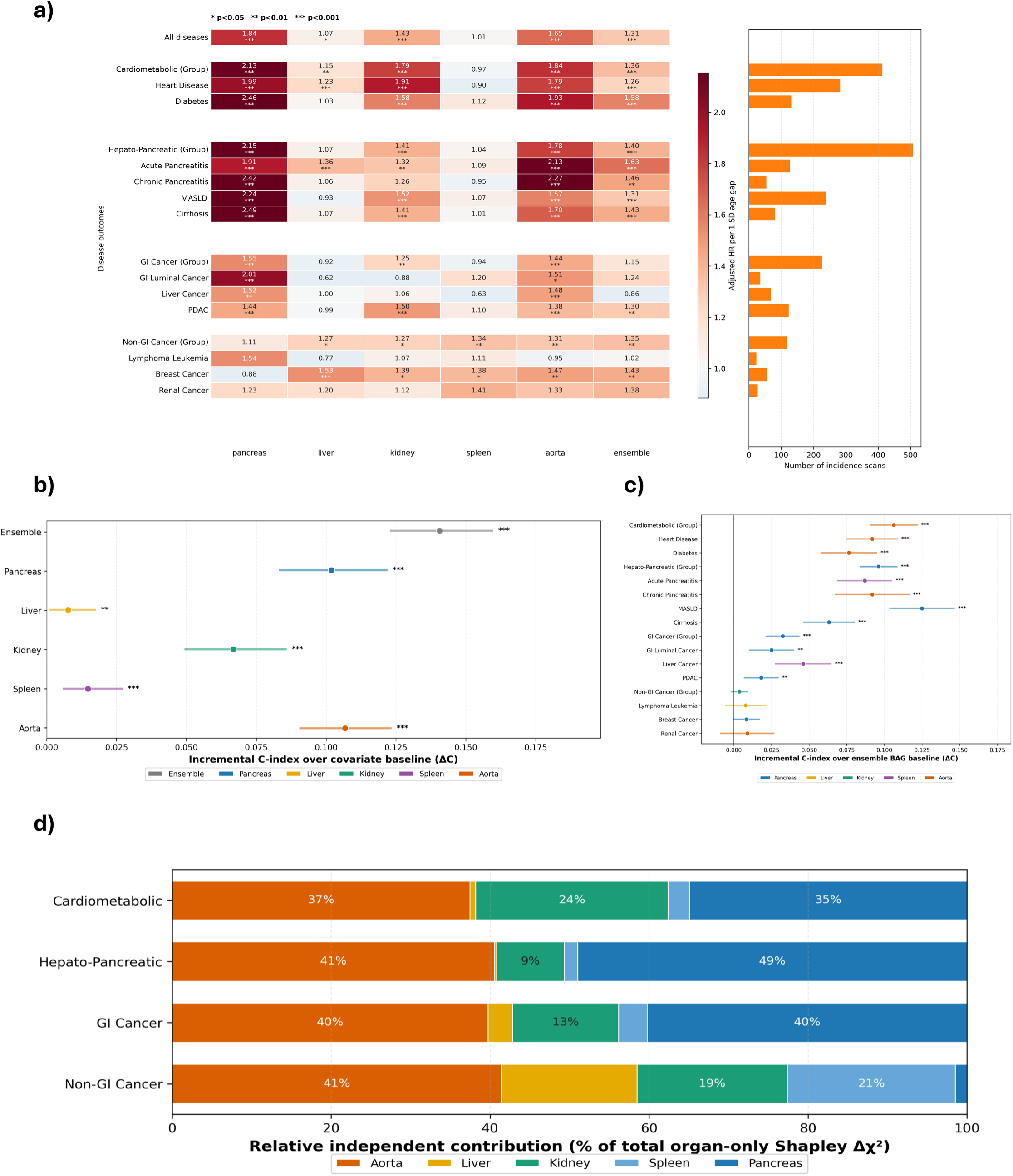
Prognostic value of organ biological age gaps (BAGs) across disease outcomes. (a) Heatmap of adjusted hazard ratios (HR) per one SD increase in organ BAG from Cox models (adjusted for chronological age, sex, and BMI), across five organ-specific BAGs and the multi-organ ensemble. Asterisks denote statistical significance (*p<0.05, **p<0.01, ***p<0.001); the right bar chart shows pre-diagnostic incidence scan counts per outcome. (b) Incremental concordance index (ΔC) of each organ and ensemble BAG over the demographic covariate baseline across all pooled outcomes; dots and lines represent point estimates and 95% bootstrap confidence intervals. (c) Per-disease ΔC of the best-performing individual organ BAG over the ensemble BAG baseline, highlighting organ-specific prognostic specificity. (d) Relative independent contributions (% of total organ-only Shapley Δχ²) per organ BAG within each disease group, derived from exact Shapley values computed over all 2⁵ = 32 Cox model subsets.

To quantify joint contributions, we applied exact Shapley value decomposition (Δχ²) across all 2⁵ = 32 Cox model subsets. Aorta BAG was the largest contributor across all four disease groups (37–41%). Within cardiometabolic diseases, kidney (24%) and pancreas (35%) provided substantial additional contributions; pancreas BAG dominated the hepato-pancreatic group (49%); and aorta and pancreas contributed approximately equally to GI cancers (∼40% each). Non-GI cancers showed a more evenly distributed profile, suggesting systemic rather than organ-specific aging as the primary driver of risk in this category.

To visualize the stratification capacity of the ensemble BAG, we stratified subjects into high-risk (BAG ≥ +1 SD) and low-risk groups and generated Kaplan–Meier cumulative incidence curves for all diseases and each major disease group (Figure 4). Across all outcomes, the high-risk group showed substantially elevated cumulative incidence relative to the low-risk group. For all diseases pooled, the Cox HR was 1.89 (95% CI: 1.68–2.13). Disease-group-specific analyses revealed particularly strong stratification for hepato-pancreatic diseases (HR=4.07 [3.37–4.91]) and cardiometabolic diseases (HR=3.44 [2.80–4.21]), with significant but more modest separation for GI cancer (HR=1.91 [1.29–2.84]) and non-GI cancer (HR=1.63 [1.12–2.36]).

**Figure 4.**
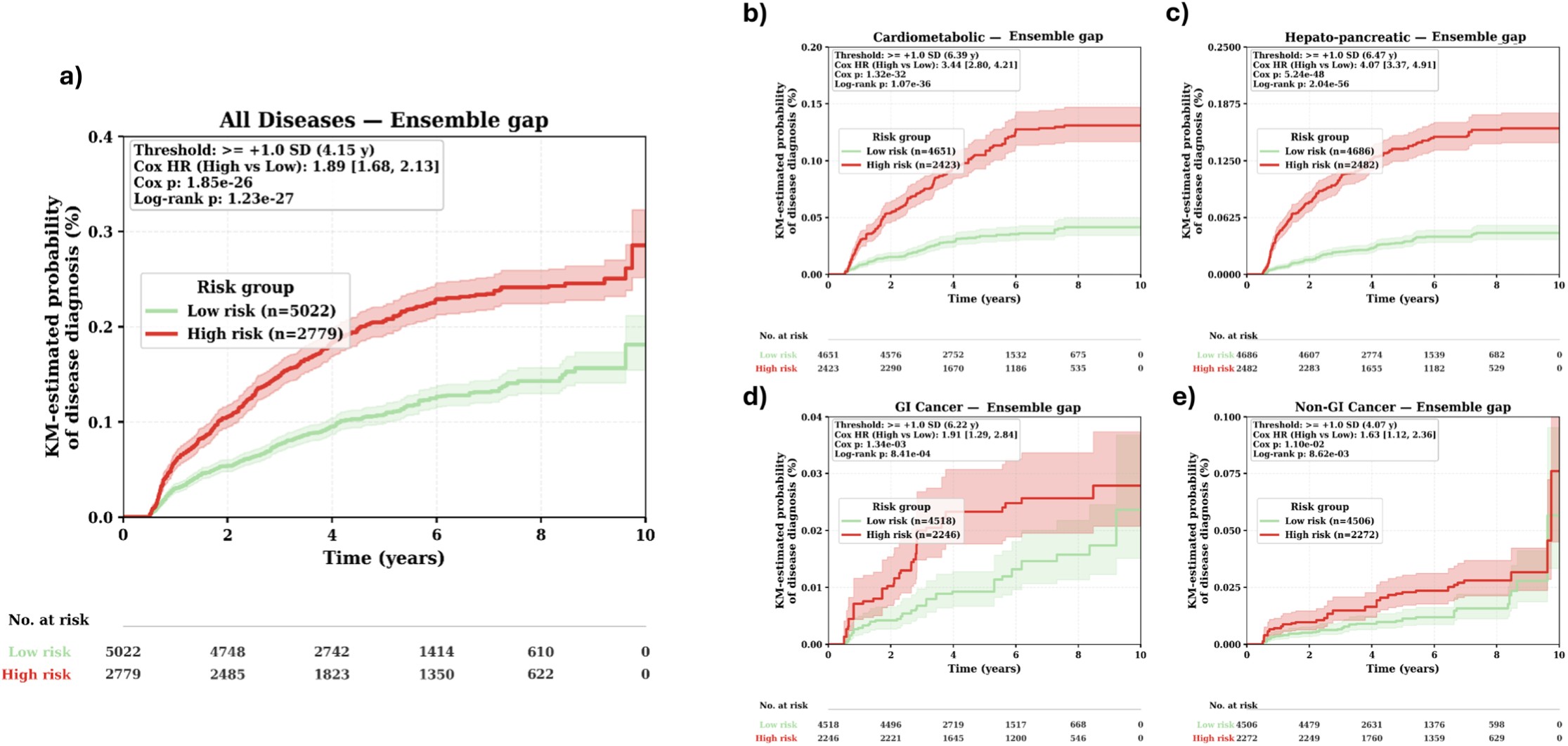
Ensemble biological age gap stratifies future disease incidence across major disease categories. Kaplan–Meier cumulative incidence curves comparing high-risk (ensemble BAG ≥ +1 SD) and low-risk subjects for (a) all diseases pooled (HR = 1.89 [1.68, 2.13]), (b) cardiometabolic diseases (HR = 3.44 [2.80, 4.21]), (c) hepato-pancreatic diseases (HR = 4.07 [3.37, 4.91]), (d) gastrointestinal cancers (HR = 1.91 [1.29, 2.84]), and (e) non-gastrointestinal cancers (HR = 1.63 [1.12, 2.36]). Cox HRs are adjusted for chronological age, sex, and BMI. The number at risk for each stratum is shown below each panel. The strongest risk separation is observed for hepato-pancreatic and cardiometabolic outcomes.

To characterize disease-associated aging patterns across abdominal organs, we examined the distributions of ensemble and organ-specific BAGs across healthy controls and the four major disease groups using ridge plots (Figure 5). Healthy controls were centered near zero across all organs, consistent with minimal age-gap deviation. In contrast, each disease group exhibited a distinct multiorgan aging signature. Cardiometabolic disease showed broad acceleration across all organs, with the strongest positive shifts in the aorta and pancreas. Hepato-pancreatic disease displayed a markedly localized pattern dominated by pancreatic aging (mean BAG >6 years), with comparatively modest shifts in other organs. GI cancer showed pronounced pancreatic acceleration alongside moderate positive shifts in the remaining organs, while non-GI cancer exhibited milder, more evenly distributed acceleration without a single dominant organ. These organ-resolved aging signatures support the notion that abdominal biological aging is spatially heterogeneous and disease-specific.

**Figure 5.**
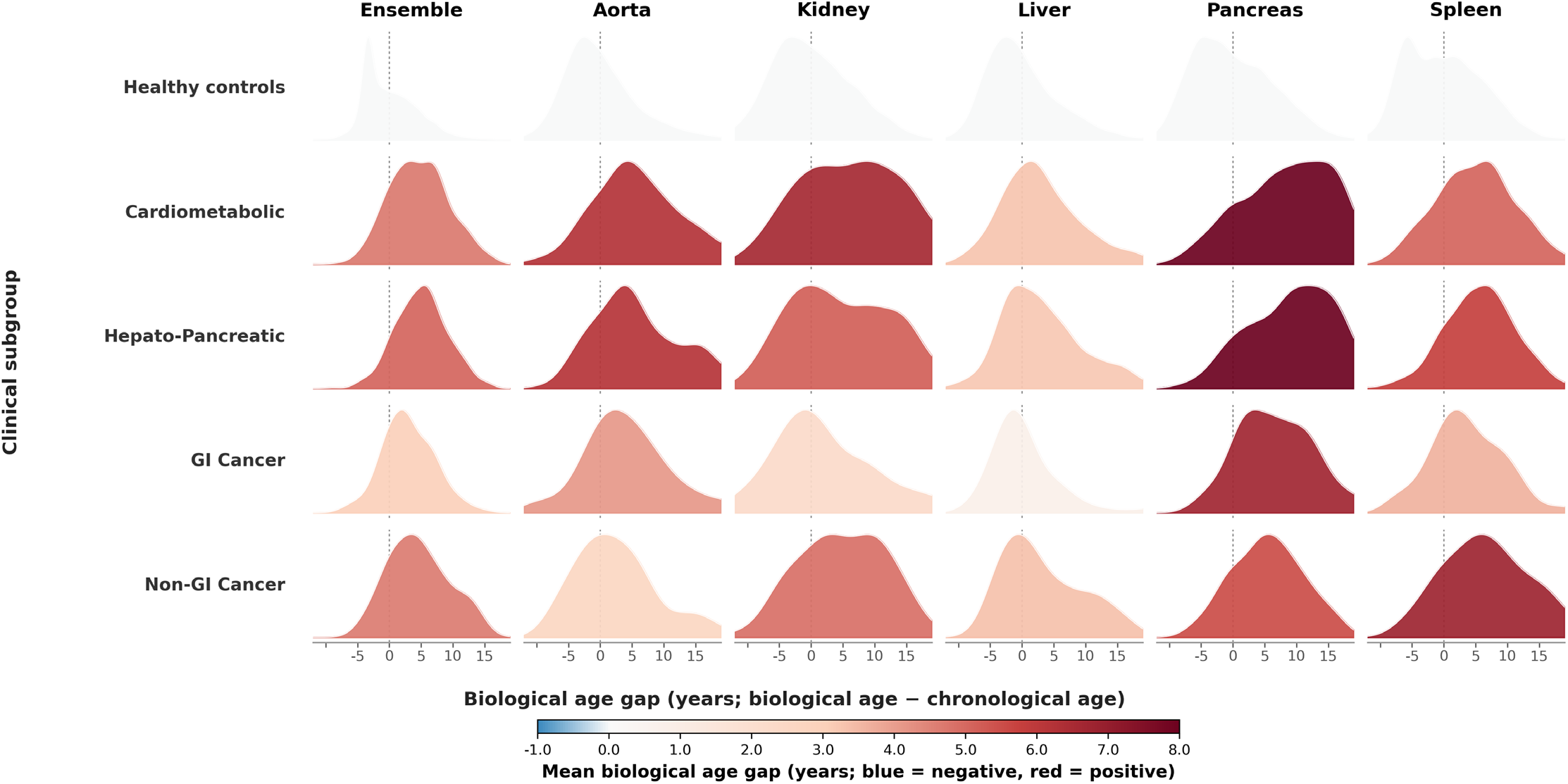
Disease-selective multiorgan aging signatures revealed by biological age gap distributions. Ridgeline plots depicting the distribution of biological age gap (BAG) for the ensemble estimate and for each organ-specific model (aorta, kidney, liver, pancreas, and spleen) across healthy controls and four major disease subgroups (cardiometabolic, hepato-pancreatic, gastrointestinal cancer, and non-gastrointestinal cancer). The dashed vertical line marks zero BAG (i.e., no deviation from expected chronological aging); fill color reflects the subgroup mean BAG, ranging from negative (blue, younger-than-expected) to positive (red, older-than-expected). Healthy controls are centered near zero across all organ models. In contrast, each disease group exhibits a distinct multiorgan aging profile: cardiometabolic disease shows broad acceleration across organs with the strongest shifts in aorta and pancreas; hepato-pancreatic disease is dominated by markedly elevated pancreatic BAG; gastrointestinal cancer shows pronounced pancreatic acceleration alongside moderate shifts in remaining organs; and non-gastrointestinal cancer displays a milder, more systemically distributed pattern without a single dominant organ.

### Ensemble biological age predicts all-cause mortality in the healthy reference cohort

To evaluate the prognostic value of our framework for all-cause mortality, we trained Cox models exclusively on the healthy control cohort to avoid disease-induced confounding. Ensemble BAG stratified subjects into significantly divergent survival trajectories (HR=1.39 [1.12–1.73], log-rank p=3.04×10⁻³; Figure 6a). When evaluated against a demographic covariate baseline (age, sex, BMI), aorta BAG achieved the highest ΔC-index (ΔC≈0.12, p<0.001), substantially exceeding that of the ensemble BAG (ΔC≈0.05, p<0.001), while pancreas and liver BAGs also contributed significantly (Figure 6b). Kidney and spleen BAGs did not reach significance over the demographic baseline alone; however, both remained independently significant when ensemble BAG was included as an additional covariate (kidney p<0.001, spleen p<0.01; Figure 6c), indicating residual organ-specific prognostic information beyond the multi-organ composite.

**Figure 6.**
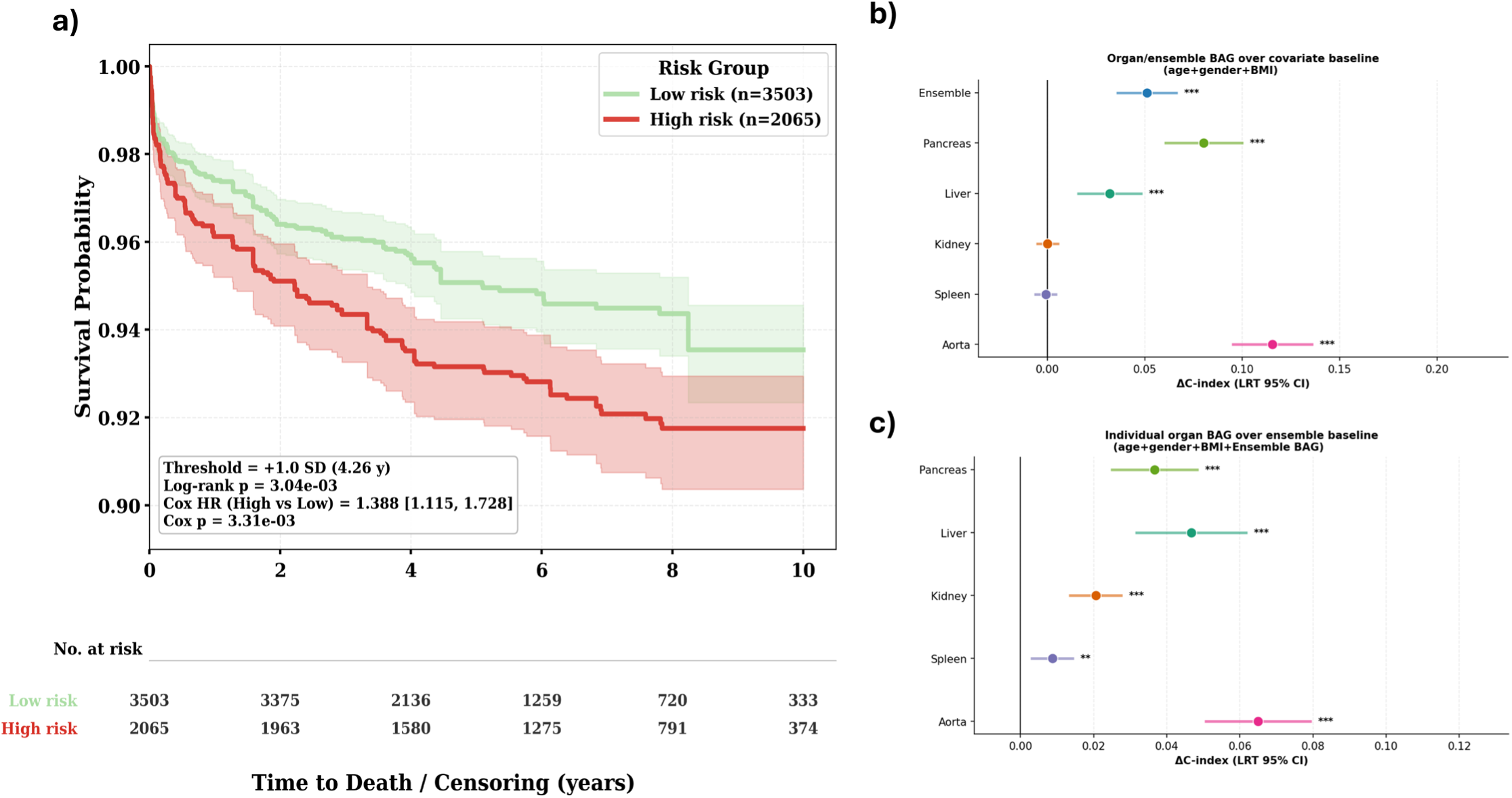
Ensemble and organ-specific biological age gaps predict all-cause mortality in a strictly healthy cohort. (a) Kaplan–Meier survival curves for all-cause mortality comparing high-risk (ensemble BAG ≥ +1 SD, n = 2065) and low-risk (n = 3503) subjects in the strictly healthy cohort (Cox HR = 1.39 [1.12, 1.73], log-rank p = 3.04×10⁻³). The number at risk for each stratum is shown below the panel. (b) Incremental concordance index (ΔC-index, 95% CI) of each organ-specific and ensemble BAG over a demographic covariate baseline (age, sex, BMI) for all-cause mortality. Aorta BAG yields the largest incremental contribution (ΔC ≈ 0.12, q < 0.001), followed by ensemble, pancreas, and liver BAGs; kidney and spleen BAGs do not reach significance against the demographic baseline alone. (c) Incremental ΔC-index of each individual organ BAG over an ensemble-augmented baseline (demographic covariates plus ensemble BAG). All five organ BAGs retain significant incremental prognostic contributions beyond the multi-organ composite, with aorta, pancreas, and liver showing the largest increments.

### Organ-specific biological age gaps align with organ-specific clinical phenotypes supporting biological plausibility

To assess the biological plausibility and clinical relevance of organ BAGs, we computed covariate-adjusted partial correlations between each BAG and a panel of laboratory measurements in the healthy cohort (Figure 7a). Across all BAG models, accelerated aging was positively associated with glucose, creatinine, hepatic enzyme markers (ALT, AST, ALP), and insulin use, and negatively associated with red blood cell (RBC) count. Organ-specific associations were also evident: liver and kidney BAGs showed the strongest correlations with hepatic enzymes (ALT, AST) and creatinine respectively, while aorta BAG and pancreas BAG were prominently associated with ALP and insulin use, consistent with their respective disease relevance.

**Figure 7.**
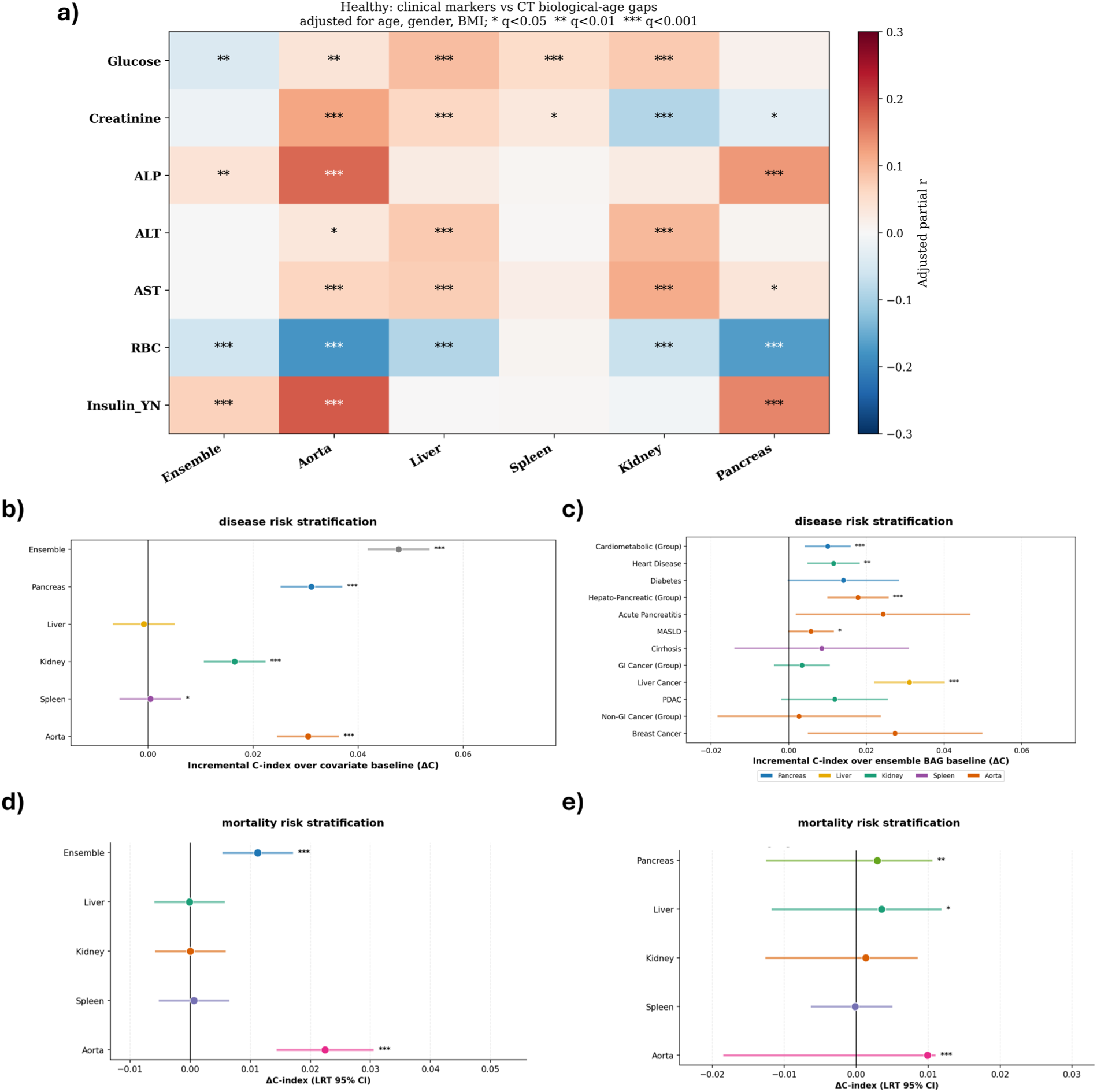
Organ-specific biological age gaps correlate with organ-relevant clinical markers and retain incremental prognostic value beyond routine blood biomarkers. (a) Heatmap of covariate-adjusted partial correlations (adjusted for age, sex, and BMI) between seven clinical laboratory variables (glucose, creatinine, ALP, ALT, AST, RBC, and insulin use) and six BAG models (ensemble, aorta, liver, spleen, kidney, pancreas) in the healthy cohort. Cell color indicates the direction and magnitude of the adjusted partial correlation coefficient (red = positive, blue = negative; scale −0.3 to +0.3); asterisks denote FDR-corrected significance. Organ-specific associations are clinically coherent: liver BAG shows the strongest positive correlations with hepatic enzymes (ALT, AST); kidney BAG with creatinine; aorta and pancreas BAGs with ALP and insulin use; and RBC is negatively associated across multiple models. (b) ΔC-index of each organ-specific and ensemble BAG over a mortality covariate baseline for all-disease risk stratification, with blood biomarkers (glucose, creatinine, RBC, ALP) added to the baseline. Ensemble, pancreas, kidney, and aorta BAGs retain significant incremental prognostic value. (c) Per-disease ΔC-index of the best-performing organ BAG over the ensemble-plus-biomarker baseline for individual disease outcomes; color denotes the contributing organ. (d) ΔC-index of organ-specific and ensemble BAGs over the biomarker-augmented baseline (age, sex, BMI, glucose, creatinine, RBC, ALP) for all-cause mortality; only ensemble and aorta BAGs retain significant incremental prognostic value. (e) ΔC-index of individual organ BAGs over the ensemble-plus-biomarker baseline for all-cause mortality; pancreas, liver, and aorta BAGs show significant residual contributions beyond both the multi-organ composite and routine laboratory markers.

To quantify the incremental prognostic value of organ BAGs beyond routine blood biomarkers (glucose, creatinine, RBC, ALP), we repeated the ΔC-index analysis with biomarkers included in the baseline model (Figure 7b). The ensemble BAG retained significant incremental prognostic value over the biomarker-augmented baseline for all-disease risk stratification (ΔC≈0.048, p<0.001). At the disease-specific level, the pattern of best-performing organ BAGs shifted relative to the biomarker-naïve analysis: liver BAG emerged as the leading predictor for liver cancer (ΔC≈0.041), and kidney BAG showed the highest increment for heart disease (ΔC≈0.012), reflecting complementary information captured by imaging beyond that encoded in blood tests. For mortality, only aorta BAG retained observable incremental prognostic value over the biomarker-augmented baseline (ΔC≈0.02–0.03), while other organ BAGs showed marginal or non-significant contributions, suggesting that the aorta’s structural aging signal is not fully captured by standard laboratory markers.

## Discussion

We developed a CT-derived abdominal biological aging framework anchored to a large strictly healthy normative cohort and evaluated its prognostic architecture through four sequential nested analyses. The principal finding is that ensemble biological age gap (BAG) — aggregating radiomic aging signals from five abdominal organs into a single composite estimate — provides substantial incremental prognostic value for future disease incidence and all-cause mortality beyond both demographic covariates and routine blood biomarkers, demonstrating that CT-derived structural aging adds to the risk information contained in standard clinical data. Within this hierarchy, organ-specific BAGs provide a second tier of prognostic specificity, selectively augmenting the ensemble estimate for anatomically focal diseases — particularly cardiovascular and hepato-pancreatic outcomes — while the ensemble estimate remains the more robust predictor for systemic and extra-abdominal malignancies. Together, these results define CT-derived abdominal biological aging as a hierarchically organized multi-level biomarker framework with the potential to enrich routine clinical risk stratification at both population and organ-resolved levels.

### Normative abdominal aging from routine CT anchors organ-wise biological aging

Because biological age cannot be observed directly, its interpretation depends on how well the model is anchored to normative aging rather than to occult or overt disease-related change. In this study, we addressed this by training and validating the abdominal age framework in a strictly curated healthy cohort spanning a broad adult age range from 20 to 100 years old. The resulting model showed close alignment with chronological age in healthy individuals across nested cross-validation, internal testing, and external validation, supporting that the learned signal reflects physiological aging patterns rather than nonspecific pathology or dataset bias. This normative anchoring is important for downstream interpretation: it strengthens the inference that positive biological age gaps observed in pre-diagnostic disease cohorts represent deviation from healthy abdominal aging, rather than residual confounding from mixed-population model development. Accordingly, the value of the healthy reference is not only improved age-prediction performance, but also a more credible basis for interpreting abdominal biological age gap as a clinically meaningful marker of accelerated aging.

### Ensemble biological age gap provides incremental prognostic value beyond demographic data and routine blood biomarkers

Establishing that a novel biomarker adds incremental prognostic value beyond available clinical information is a necessary precondition for clinical translation. In sequential nested Cox analyses (Analysis A1), the ensemble BAG improved concordance for future disease incidence by ΔC=0.141 beyond a demographic covariate baseline of chronological age, sex, and BMI — a substantial increment that was consistent across all four major disease categories. For all-cause mortality in strictly healthy subjects, ensemble BAG similarly improved concordance by ΔC≈0.051 over the demographic baseline. Kaplan–Meier stratification by ensemble BAG demonstrated practically significant risk separation: the high-risk group (≥+1 SD) showed substantially elevated cumulative disease incidence (HR=1.89, 95% CI: 1.68–2.13, p=1.85×10⁻²⁶) and higher all-cause mortality in healthy subjects (HR=1.39, 95% CI: 1.12–1.73, p=3.04×10⁻³, Figure 6a), supporting the utility of ensemble BAG as a broad-spectrum risk stratification signal across both disease incidence and survival.

A central question in biological aging biomarker development is whether a new measure adds information beyond what clinicians already obtain from routine clinical tests [39]. When fasting glucose, creatinine, red blood cell count, and alkaline phosphatase were included in the baseline model (Analysis A3), ensemble BAG retained significant incremental prognostic value for future disease incidence (ΔC=0.048, p<0.001), demonstrating that CT-derived structural aging adds prognostic information beyond what systemic laboratory measurements alone provide. Conceptually, blood-based biomarkers largely reflect circulating functional states — metabolic regulation, renal filtration, hepatic enzyme activity, and hematopoiesis — whereas CT-derived radiomic features capture tissue-level structural changes in the form of organ texture, attenuation heterogeneity, volumetric remodeling, and morphological variation that are not fully mirrored by any conventional laboratory panel. This is analogous to the way structural brain MRI–derived age gaps provide prognostic information beyond neurological blood markers [5,6], and consistent with prior multi-organ molecular aging studies showing that organ-resolved biological age clocks capture disease-relevant variation complementary to circulating biomarkers [11,38]. At the disease-specific level, the pattern of leading organ predictors shifted when moving from the biomarker-naïve to the biomarker-augmented baseline: liver BAG emerged as the leading predictor for liver cancer (ΔC≈0.041) and kidney BAG for heart disease (ΔC≈0.012), reflecting organ-specific information overlap between imaging and laboratory domains. For all-cause mortality, only aorta BAG retained meaningful incremental prognostic value beyond the biomarker-augmented baseline (ΔC≈0.02–0.03), consistent with the limited ability of standard laboratory panels to capture vascular structural aging accessible through direct CT assessment of the aortic wall [^18^]. Taken together, ensemble and organ-specific BAGs function as complementary rather than redundant biomarkers relative to routine blood tests, supporting the view that CT-derived structural aging provides an incrementally informative layer within the clinical biomarker ecosystem.

### Organ-specific biological age gaps reveal hierarchically organized, disease-selective aging beyond ensemble

A second key question is whether maintaining organ-level decomposition provides prognostic utility beyond the ensemble estimate, or whether a single composite is sufficient. Per-disease incremental analyses (Analysis A2) demonstrated that individual organ BAGs added statistically significant concordance beyond the ensemble-augmented baseline for anatomically focal diseases: aorta BAG yielded the largest incremental improvement for cardiovascular outcomes (ΔC=0.091), and pancreas BAG provided the highest increment for hepato-pancreatic disease. This pattern is consistent with the principle established in multi-organ proteomic and molecular aging literature that organ-specific biological age clocks exhibit disease-selective associations, with each organ biomarker most informative for diseases that primarily involve that organ system [11,38]. The present data extend this principle from the molecular to the CT-derived structural domain and demonstrate that the same hierarchical organization holds: ensemble aggregation improves robustness and breadth, while organ-level resolution adds specificity for focal disease.

Exact Shapley value decomposition across all 2⁵=32 organ subsets provided quantitative support for this hierarchical structure. Aorta BAG was the largest independent contributor across all four disease groups (37–41%), reflecting broad vascular aging relevance irrespective of disease category. Within disease-specific groupings, pancreas BAG dominated the hepato-pancreatic group (49%), while kidney (24%) and pancreas (35%) provided substantial secondary contributions within cardiometabolic disease. Non-GI cancers exhibited a more evenly distributed organ contribution profile, consistent with systemic rather than organ-localized aging as the primary driver, and correspondingly showed the weakest incremental organ BAG signal over the ensemble in per-disease ΔC analyses. This supports the clinical principle that organ decomposition is most informative for diseases with anatomically localized pathophysiology, while the ensemble estimate performs more reliably when disease biology is diffuse or arises from extra-abdominal sites. For all-cause mortality, aorta BAG was the single strongest predictor (ΔC≈0.12 over demographic baseline), substantially exceeding the ensemble BAG (ΔC≈0.05) in this outcome-specific analysis, suggesting that the relative advantage of organ-specific over ensemble BAG is itself outcome-dependent — and most pronounced when vascular structural aging is the dominant mortality pathway. The disease-selective organ aging signatures observed in ridgeline plots further corroborated this organization: each disease group exhibited a distinct multi-organ BAG profile, with hepato-pancreatic disease showing a markedly localized pancreatic aging pattern (mean BAG >6 years), cardiometabolic disease exhibiting broad acceleration with the largest shifts in aorta and pancreas, and non-GI cancer displaying milder, more evenly distributed acceleration without a dominant organ. This spatial heterogeneity of abdominal aging across disease categories is consistent with organ-selective molecular aging signatures reported in proteomic aging studies [38].

Cross-organ coupling further modulates the organ-specific signal in a disease-selective manner. For cirrhosis, the liver BAG did not rank as the single strongest predictor over the ensemble baseline, yet it retained a significant HR (1.41) in hazard ratio analyses — an observation consistent with hepato-pancreatic co-vulnerability, where early cirrhosis-associated structural change may be reflected across multiple tightly coupled abdominal organs rather than exclusively in the liver. In contrast, for acute pancreatitis, pancreatic BAG was the dominant predictor over ensemble, suggesting that the pancreatic structural aging signal is sufficiently organ-specific to provide incremental information even after accounting for the composite multi-organ estimate. Finally, organ-specific BAGs retained incremental prognostic contributions beyond a combined ensemble-plus-biomarker baseline (Analysis A4) in targeted pairings — liver BAG for liver cancer and kidney BAG for heart disease — confirming that organ-specific structural aging information is additive across all four analytical tiers: demographic covariates, ensemble BAG, blood biomarkers, and organ-specific BAG. This four-level sequential hierarchy supports the potential value of organ-resolved CT-derived aging for clinical scenarios in which targeted organ risk assessment is warranted beyond general systemic screening.

### Biomarker analysis links CT age gaps to organ physiology

Clinical correlations and radiomic patterns support biological plausibility. The associations with laboratory and diagnostic variables support the biological plausibility of BAG while remaining inferential rather than mechanistic. Rather than showing diffuse or nonspecific correlations, organ-specific and ensemble age gaps aligned with clinically coherent markers of metabolic, hepatic, renal, and vascular dysfunction. This pattern is consistent with prior CT literature showing age-related changes in abdominal organ phenotype, including altered liver and pancreatic attenuation, age-dependent liver volume variation, increasing pancreatic fat, and later-life decline in pancreatic parenchymal volume [^19–23^].

Across organs, the retained radiomic signature was enriched for biologically interpretable feature families, including first-order intensity distribution, gray-level non-uniformity and variance, neighborhood coarseness or busyness, run-length and size-zone emphasis, and shape or volume descriptors. In the liver and spleen, these patterns are concordant with CT evidence that fibrosis severity is reflected by hepatosplenic volumetric remodeling and that combined hepatic-splenic radiomic signatures improve fibrosis stratification [^24,25^]. In the pancreas, the association of older age gaps with insulin use, HbA1c, glucose, and BMI is biologically consistent with age- and metabolic-associated pancreatic fatty infiltration, age- and sex-related variation in pancreatic CT attenuation, and decline in pancreatic volume in later life; prior CT radiomics studies further show that pancreatic texture features can capture chronic structural injury even when conventional imaging findings are subtle, supporting the interpretability, though not direct mechanistic specificity, of pancreas-derived BAG signals [^21–23,26^]. In the kidney, the relationship between kidney age gap and creatinine is similarly plausible given CT evidence that renal parenchymal thickness decreases with age, together with broader aging biology implicating nephrosclerosis, tubular atrophy, and interstitial fibrosis, and recent work linking CT radiomic features to chronic histopathologic injury on native kidney biopsy [^27–29^]. In the aorta, the observed signal is likewise credible, as abdominal aortic calcium burden and aortic diameter both increase with age on CT, suggesting a structural substrate for variation in aortic radiomic and shape-derived features [^30,31^].

Higher RBC was associated with younger age gaps across several models, a direction broadly consistent with hematologic literature showing that RBC-related measures increase to adulthood and tend to decline with advanced age, particularly in men, while anemia prevalence rises in older adults [^32–34^]. Because age effects on RBC indices are sex- and cohort-dependent, this association should be interpreted as supportive rather than definitive. Although most correlations were modest, this is expected for a composite aging biomarker that captures tissue-level variation not represented by any single clinical variable. Taken together, these findings suggest that BAG reflects both shared systemic aging processes and organ-specific vulnerability, supporting the biological plausibility and clinical interpretability of the framework for risk stratification.

### Limitations

This study has several limitations. First, the model-development cohort was retrospectively assembled from clinically indicated abdominal CT examinations performed for any indication at a single tertiary-care center between 2012 and 2022. As such, the cohort is not population-based and is likely enriched for comorbidity, diagnostic evaluation, surveillance imaging, and other healthcare-utilization patterns that may influence both radiomic features and downstream disease risk. Although external validation in multi-center healthy cohorts supports the transportability of age prediction, and external pancreas biological age analyses showed discrimination between PDAC and non-PDAC CT datasets, broader validation of disease-association and disease-prediction performance across independent health systems remains necessary.

Second, outcome ascertainment relied on ICD codes and EHR radiology reports, which are susceptible to incomplete capture, documentation variability, and phenotyping misclassification. Third, association analyses were adjusted only for sex and BMI, leaving open the possibility of residual confounding by additional clinical, behavioral, and sociodemographic factors. Fourth, because the model was developed from routine clinical CT examinations acquired over a decade and across both contrast-enhanced and non-contrast protocols, heterogeneity in scanner hardware, acquisition parameters, reconstruction settings, and contrast administration may have influenced radiomic measurements. While this design improves robustness to real-world imaging variation, it may also reduce sensitivity to phase-specific radiomic information relevant to biological aging.

Because the strongest reported prognostic analyses focus on adults aged 20–60 years, effect sizes should not be generalized directly to older adults. A sensitivity analysis in adults aged 20–80 years (Supplementary Figure S3) showed persistence of several organ-specific signals, particularly pancreas and aorta BAG, but attenuation of other biomarkers, consistent with age-dependent modification of relative risk.

**In conclusion,** we have established a hierarchically organized, CT-derived abdominal biological aging framework in which ensemble BAG provides broad-spectrum prognostic value beyond both demographic covariates and routine blood biomarkers, and organ-specific BAGs provide a second tier of disease-selective prognostic specificity beyond the ensemble estimate. Sequential incremental analyses confirm that CT-derived structural aging provides incremental prognostic information beyond conventional clinical data — whether that baseline is demographic alone or augmented with laboratory markers — and that organ decomposition adds clinical value specifically for anatomically focal diseases such as cardiovascular and hepato-pancreatic conditions. These findings establish CT-derived abdominal biological aging as a multi-level biomarker hierarchy with potential for clinical translation in risk stratification, targeted organ surveillance, and early disease detection. Future studies should evaluate this framework in prospective multi-center cohorts, assess performance across contrast-protocol strata, and explore protocol-aware, organ-specific deep learning extensions to further improve biological age estimation and disease-specific predictive utility.

## Methods

In this study, we proposed three hypotheses:

1. There is a negligible difference between abdominal biological age and chronological age on healthy subjects with only aging related deformation. Such assumption justifies using chronological age as an estimate of ground truth biological age if the model is trained solely on healthy cohorts.
2. Biological age will deviate from chronological age during development of diseases, which therefore can serve as a biomarker for disease prediction.
3. The individual-organ biological age can deviate from the abdominal biological age, reflecting the specific health conditions of each organ. In addition, accelerated biological aging varies across different organs, indicative of the relevance of each organ towards the disease.

### Data Collection and Description

All datasets included in this study were approved by the institutional review board (IRB) of the participating medical center. All scans of the subjects are anonymized prior to the downloading for compliance with privacy regulation.

The main cohort comprised 21,759 healthy abdominal CT scans from 11,917 subjects aged 20–100 years, acquired between 2012 and 2022. Healthy scans were defined by the absence of ICD-codes within EHR related to cancer and common abdominal diseases, including cirrhosis and pancreatitis, as well as major extra-abdominal diseases relevant to this study, such as ischemic heart disease. The main cohort is split into 12,737 scans from 6,293 subjects for training and validation, and 9,022 scans from 5,624 subjects as internal independent test set. We additionally assembled a disease cohort of 44,844 CT scans from 18,900 subjects with at least one post-diagnosis CT scan and, where available, pre-diagnostic CT scans obtained up to 10 years before diagnosis. For external validation, we included two independent datasets: the PANORAMA dataset, consisting of 1,959 venous-phase contrast-enhanced CT scans with PDAC and non-PDAC labels, and a multi-center healthy dataset of 120 strictly healthy CT scans. These external datasets were used to assess biological age gap associations with PDAC and the transferability of age prediction to unseen healthy populations, respectively. Detailed demographic information for all datasets is provided in **Table S1**.

### Imaging Acquisition and Segmentation

All CT scans from the subjects were anonymized and downloaded in DICOM format and then converted to NIFTI format. In addition, image resampling, Hounsfield Unit windowing, and intensity standardization are applied to harmonize all CT scans to control data heterogeneity. An initial multi-organ segmentation was performed using a robust nnUNet model trained with domain randomization [^35^]. Segmentation outputs underwent automated radiomics based quality control, with visual review and manual refinement performed by local radiologists for scans flagged by dropping below a confidence threshold.

### Radiomic Feature Extraction and Selection

For our study, six organs, specifically pancreas, liver, spleen, abdominal aorta, left and right kidney, were selected due to the close relation to whole body physiology and relatively obvious change through aging. Features used in this study were extracted using PyRadiomics. Common image filters and radiomics groups were adopted for radiomics extraction to exhaustively search for potential shape-based and texture-based features related to aging. Extracted features are standardized to minimize data heterogeneity. Additionally, we merged features from left and right kidney into a single feature group called kidney by averaging each type of feature within two groups.

For each selected organ, a three-stage feature selection pipeline was implemented to filter out irrelevant and duplicated features. First, age-to-feature Pearson Correlation Coefficient (PCC) was computed on all features within the feature group and irrelevant features with PCC lower than 0.35 were excluded. Second, feature-to-feature PCC is applied across filtered features, and two features with PCC larger than 0.60 were processed so that the one with highest age-to-feature PCC is preserved. Third, if the filtered features were still more than 10 features, we apply the recursive feature elimination with LASSO Regression as the estimator to recursively eliminate least relevant features to reach the desired feature number. To additionally constraint on duplicate features from same image filter groups, we also constrain the maximum number of same radiomics feature within the same image filter group (wavelet, LoG, etc.) to be 2.

### Model Development

We adopted a two stage ensembled machine learning network with XGBoost as the backbone. For each organ, a separate XGBoost model was trained to generate organ-specific biological age, and an ensemble XGBoost model was trained on the organ-specific biological ages generated from organ-level age prediction models. We also tested the performance with other common ML models like SVR, LASSO Regression, Random Forest Regression and Multi-Layer Perceptron, and XGBoost achieves highest performance and was therefore adopted as the backbone of whole model.

To robustly verify the performance of our biological age prediction model, we followed the work done by Goallec et al. [9] to implement nested cross validation with stratification on age and gender was implemented during the training process. For each organ-level age prediction model, we performed separate feature selection and model training on each fold of the inner cross-validation, and the model with the best validation performance was selected for evaluation on the outer test fold.

Similarly, the ensembled abdominal age model was trained and validated on the same inner fold and tested on the same outer test fold, using the predicted age of corresponding organ-level model.

### Downstream cohorts and biological age-gap phenotypes

As mentioned in many prior works, regression model for age prediction naturally possesses the issue of biasing extreme values towards the mean value [^10,11,36^]. Therefore, we fitted a simple linear model on the validation dataset of each corresponding organ level and ensemble model so that given the CA, the model will output an offset value to correct the extreme value bias. Noted that this bias correction is only applied during evaluation that are not accuracy related to avoid data leakage issue.

For downstream analyses, organ-specific biological ages were generated for the aorta, kidney, liver, pancreas and spleen, and were further combined to obtain an ensemble abdominal biological age. Biological age gap (BAG) was defined as predicted biological age minus chronological age for each organ-specific model and for the ensemble model. Because age-prediction models can retain residual dependence on chronological age, downstream analyses were performed using bias-corrected BAG phenotypes. When direct comparison across organs was required, BAG values were further standardized to place biomarkers on a common scale.

The primary downstream disease analyses used scans from the healthy control cohort together with pre-diagnostic scans from subjects who subsequently developed disease. To focus on early disease-associated deviation from normative aging, only scans acquired 6 months to 10 years before the first recorded diagnosis were included in the time-to-event analyses. Based on EHR and ICD-derived phenotyping, outcomes were grouped into four major categories: cardiometabolic disease, hepato-pancreatic disease, gastrointestinal cancers and non-gastrointestinal cancers. For descriptive analyses of disease-selective organ aging patterns, the same organ-specific and ensemble BAG phenotypes were evaluated across the healthy cohort and the major disease subgroups. This framework enabled all downstream analyses to be anchored to the same normative age-prediction model while testing how organ-resolved deviation from expected aging related to future disease.

Because ensemble BAG associations with disease risk showed marked attenuation with advancing chronological age, reaching near-null by the early 70s (Supplementary Figure S2), all primary prognostic analyses — Cox models, ΔC-index comparisons, Kaplan–Meier stratification, and Shapley decomposition — were restricted to subjects aged 20–60 years. The 20–60-year range was selected after age-interaction analyses showed attenuation of relative BAG-associated risk with advancing chronological age. These focused analyses are therefore interpreted as prevention-oriented subgroup analyses rather than as evidence that BAG lacks utility in older adults.

### Risk stratification and organ-resolved disease analysis

Associations between biological age gaps and future disease were evaluated using Cox proportional hazards models adjusted for chronological age, sex, and body mass index (BMI). Separate models were fitted for each organ-specific and ensemble BAG across all disease outcomes. Hazard ratios and confidence intervals were estimated per 1-SD increment in BAG. For each disease outcome, the leading organ-specific BAG was identified as the predictor whose full model yielded the largest incremental likelihood-ratio chi-square statistic (Δχ²) over the demographic covariate baseline (Figure 3a).

To quantify incremental prognostic contributions beyond established clinical baselines, four sequential nested Cox analyses (A1–A4) were applied across two outcome domains — future disease incidence and all-cause mortality. In each nested comparison, a baseline Cox model was fitted first and the BAG of interest was added as an additional predictor; the ΔC-index (difference in Harrell’s concordance index between the full and baseline models) quantified incremental prognostic contribution.

**A1 —** each BAG (five organ-specific plus ensemble) vs. a demographic covariate baseline (chronological age, sex, BMI), establishing marginal prognostic contributions above routinely available clinical information (Figures 3b and 6b);

**A2 —** each individual organ BAG vs. an ensemble-augmented baseline (demographic covariates plus ensemble BAG), isolating residual organ-specific information beyond the multi-organ composite; for disease outcomes the organ with the highest ΔC-index over this baseline is reported per disease category (Figure 3c); for mortality all five organ BAGs are reported (Figure 6c);

**A3 —** each BAG (five organ-specific plus ensemble) vs. a biomarker-augmented baseline (demographic covariates plus fasting glucose, creatinine, red blood cell count, and alkaline phosphatase), quantifying incremental prognostic value beyond routine laboratory markers (Figures 7b and 7d);

**A4 —** each individual organ BAG vs. an ensemble-plus-biomarker baseline (demographic covariates, routine blood biomarkers, and ensemble BAG), identifying residual organ-specific contributions beyond both the composite aging signal and standard laboratory information; for disease outcomes the best organ per disease category is reported (Figure 7c); for mortality all five individual organs are reported (Figure 7e).

The ΔC-index point estimate was computed from the full dataset for each comparison. For disease-pooled analyses (A1 and A3 for disease outcomes), 95% confidence intervals were derived using event-stratified bootstrap resampling (300 iterations), resampling case and control subjects separately to preserve event rates; significance thresholds were set at p < 0.05, p < 0.01, and p < 0.001.

To quantify joint independent contributions of organ BAGs within each major disease group, exact Shapley value decomposition was applied across the five organ-specific BAGs (excluding ensemble). Cox proportional hazards models were fitted for all 2^5^ = 32 possible organ subsets, with demographic covariates retained in each model. Disease groups for which any of the 32 subset Cox fits failed convergence were excluded.

To visualize ensemble BAG stratification capacity, subjects were divided into high-risk (BAG ≥ +1 SD) and low-risk groups. Kaplan–Meier cumulative incidence curves were generated for each major disease category and for all-cause mortality in the strictly healthy cohort, with log-rank p-values and adjusted Cox hazard ratios (95% CI) annotated per stratum comparison (Figures 4 and 6a).

Distributions of ensemble and organ-specific BAG values across the healthy control cohort and major disease subgroups were visualized with ridgeline plots, with fill color reflecting subgroup mean BAG and a reference line at zero deviation (Figure 5).

### Clinical correlation and spatial interpretability analysis

To assess biological plausibility, covariate-adjusted partial correlations (adjusted for chronological age, sex, and BMI) were computed between each organ-specific and ensemble BAG and a panel of seven clinical laboratory variables — glucose, creatinine, alkaline phosphatase (ALP), alanine aminotransferase (ALT), aspartate aminotransferase (AST), red blood cell (RBC) count, and insulin use — in the strictly healthy cohort. Results were summarized as a correlation heatmap in which cell color encodes the direction and magnitude of the adjusted partial correlation coefficient (Figure 7a).

### Ethics

This study was approved by the Institutional Review Board of Cedars-Sinai Medical Center (Protocol No. Pro00053107). All data used were anonymized prior to analysis, and informed consent was waived in accordance with the IRB approval.

### Statistical analysis and reproducibility

No formal sample-size calculation was performed; cohort size was determined by the availability of de-identified abdominal CT examinations and linked electronic health record data meeting predefined cohort, image-quality, organ-availability, and analysis-specific eligibility criteria. Age-prediction models were developed using the healthy reference cohort only. Feature selection, model training, and bias correction were performed within the training/validation framework before application to internal test, external validation, disease, and mortality cohorts. Biological age gaps were bias-corrected to reduce residual chronological-age dependence and standardized when compared across organs. For downstream analyses, Cox proportional hazards models were used to estimate associations between biological age gaps and future disease or mortality outcomes, with adjustment for chronological age, sex, and body mass index unless otherwise specified. Pre-diagnostic disease analyses used CT scans obtained 6 months to 10 years before first recorded diagnosis. Kaplan–Meier curves were used for visualization, with log-rank tests and adjusted Cox hazard ratios reported for group comparisons. Proportional-hazards assumptions were assessed using Schoenfeld residuals.

Incremental prognostic value was quantified as the change in Harrell’s concordance index between nested Cox models. Confidence intervals for ΔC-index estimates were computed using subject-level bootstrap resampling, with repeated scans from the same subject resampled together when applicable. No confidence-interval truncation or post hoc narrowing was applied. All statistical tests were two-sided.

The study was retrospective and observational. Investigators were not blinded to EHR-derived cohort labels during statistical analysis. All analyses were conducted in Python 3.12; code is available from the corresponding authors upon reasonable request after publication.

### Statistical software

Organ biological age gap (BAG) predictions were generated using gradient-boosted regression trees implemented in XGBoost (v3.0.2) with scikit-learn (v1.6.1). Survival analyses were performed using Cox proportional hazards models from the lifelines library (v0.30.0); likelihood ratio tests were implemented with SciPy (v1.15.2) and statsmodels (v0.14.5). All analyses were conducted in Python

3.12 with pandas (v2.2.3) and NumPy (v1.26.4). Visualizations were generated with matplotlib (v3.10.1).

## Data Availability

All data produced in the present work are contained in the manuscript

https://panorama.grand-challenge.org/datasets-imaging-labels/

## Declarations

### Supplementary information

Supplementary Notes, Supplementary Tables S1–S2, and Supplementary Figures S1–S3 are provided in a separate Supplementary Materials file.

### Competing interests

The authors declare no competing interests Data availability

The Cedars-Sinai clinical imaging and electronic health record datasets used in this study are not publicly available because they contain patient-derived clinical imaging and health information and are subject to institutional review board approval, privacy regulations, and institutional data-use restrictions. De-identified aggregate data supporting the conclusions are included in the manuscript and supplementary materials. Individual-level CT images, radiomic features, laboratory values, diagnosis labels, and survival data cannot be publicly distributed. The PANORAMA dataset was used under its respective data-access terms, and access is governed by the original dataset providers.

### Code availability

Analysis code may be made available from the corresponding author upon reasonable request, subject to institutional approval and restrictions related to patient-derived clinical data.

