## Supplemental Materials for "Hierarchical organ aging signatures from routine abdominal CT add incremental disease risk stratification beyond blood biomarkers"

Affiliations

### Supplementary Note 1. External transportability and PANORAMA diagnostic validation

To evaluate whether the organ-wise biological age framework was sensitive to disease-related variation in an external and clinically heterogeneous setting, we performed a supplementary analysis using the PANORAMA cohort, which consists of abdominal Contrast-Enhanced CT scans with multiple levels of detection on PDAC. In this analysis, PANORAMA was treated as a clinical external cohort, not as a normative healthy validation set. Subjects were divided into two groups using the dataset PDAC label field: PAN_NONPDAC for label 0 and PAN_PDAC for label 1. During preprocessing, PANORAMA data were restricted to the required metadata and organ-radiomics columns, age was coerced to numeric format, sex/gender was harmonized, left and right kidney radiomics were averaged to generate a single kidney feature set, and subjects with missing radiomics across the required organs were excluded. Only subjects with binary labels 0 or 1 were retained.

Biological age inference was then performed with the locked organ-wise model used throughout the study. Organ-specific predictions were generated for the aorta, spleen, kidney, liver and pancreas, followed by a meta-learner that produced the abdominal ensemble prediction. In addition, a non-pancreas ensemble was constructed as the mean of the aortic, splenic, renal and hepatic predicted ages, to separate pancreas-local from broader extra-pancreatic signal. Raw biological age gap (BAG) was defined for each biomarker as predicted age minus chronological age.

Because BAG may retain chronological-age dependence, corrected BAG phenotypes were derived within each analysis window by fitting reference models in the in-house healthy cohort (IH_HEALTHY) and then residualizing and standardizing the raw BAG values. In the implemented workflow, the in-house healthy cohort served as the normative reference for this correction step. Specifically, for each BAG phenotype, a spline-based reference model was fitted as a function of age and categorical sex, and the resulting residuals were standardized to produce zBAG. The pipeline was implemented with flexible age windows (<60, all ages, <65), consistent with the broader PANORAMA analysis plan.

Two complementary statistical analyses were performed. First, a three-cohort adjusted comparison was used to quantify clinical shift and PDAC-specific excess deviation by comparing IH_HEALTHY, PAN_NONPDAC and PAN_PDAC. For each corrected biomarker, linear models were fitted with cohort as the main factor and age spline plus sex as covariates, and planned contrasts were extracted for PAN_NONPDAC − IH_HEALTHY, PAN_PDAC − PAN_NONPDAC, and PAN_PDAC − IH_HEALTHY. In the implemented code, these models were fitted with spline-adjusted age terms and heteroscedasticity-robust inference.

Second, a PANORAMA-only PDAC association analysis was performed within PAN_NONPDAC and PAN_PDAC. Logistic regression models were fitted with PDAC status as the outcome and corrected BAG as the predictor of interest, adjusting for age spline and sex. Odds ratios were reported per 1-s.d. increase in biomarkers. Primary emphasis was placed on pancreatic zBAG, non-pancreatic ensemble zBAG, and full-ensemble zBAG, while individual-organ analyses were considered exploratory. The plotted distributions in the figure correspond to pancreatic and non-pancreatic ensemble zBAG within PANORAMA, and the odds-ratio panel summarizes the PANORAMA-only logistic models.

The three-cohort comparison showed that PAN_NONPDAC already differed from IH_HEALTHY across several biomarkers, indicating that the PANORAMA non-PDAC cohort was not equivalent to a healthy baseline. Instead, the biological age framework detected measurable deviation in this clinically heterogeneous non-PDAC population relative to the in-house healthy reference. This pattern was present not only for the pancreas but also for several non-pancreatic biomarkers, consistent with sensitivity of the framework to broader disease burden and external clinical heterogeneity. This is reflected in the positive or negative adjusted PAN_NONPDAC − IH_HEALTHY shifts observed across organs in the top panel of the figure.

Against that shifted non-PDAC background, PDAC cases showed an additional and clearly organ-specific signal. The largest positive PAN_PDAC − PAN_NONPDAC difference was observed for the pancreas, whereas the corresponding effect for the non-pancreatic ensemble was small and centered near the null. The full abdominal ensemble showed weaker discrimination than pancreas-specific biological age, indicating that aggregation across organs diluted the disease-localized signal rather than strengthening it. In the lower distribution panels, pancreatic zBAG was visibly shifted upward in PAN_PDAC relative to PAN_NONPDAC, whereas the non-pancreatic ensemble remained substantially overlapping between groups.

The PANORAMA-only logistic models were concordant with these distributional results. Pancreatic zBAG showed a positive association with PDAC status, with an odds ratio above 1 per 1-s.d. increase, whereas the non-pancreatic ensemble and the full ensemble were weaker and closer to the null. Together, these findings indicate that the most reproducible disease-associated signal in PANORAMA was pancreas-local, rather than a uniformly distributed multi-organ signal.

This supplementary external analysis supports two related conclusions. First, the organ-wise biological age framework is sensitive to heterogeneous clinical disease burden in the PANORAMA non-PDAC cohort. Because PAN_NONPDAC already showed systematic deviation relative to IH_HEALTHY, these external non-PDAC subjects should not be interpreted as a normative control group. Rather, the observed PAN_NONPDAC − IH_HEALTHY shift indicates that biological age outputs respond to clinically enriched non-PDAC pathology and related cohort effects. In that sense, the framework is disease-sensitive even outside the PDAC subgroup, which is consistent with the intended biological-aging interpretation rather than a purely pancreas-cancer detector.

Second, within PANORAMA, this disease sensitivity is organ specific. PDAC cases showed additional pancreatic age-gap elevation beyond the already shifted non-PDAC clinical background, while non-pancreatic and full-ensemble effects were weaker. This pattern supports the value of maintaining organ-wise biological age decomposition: the pancreas-derived biomarker preserved local disease sensitivity, whereas global aggregation partially obscured that signal. The external PANORAMA analysis therefore validates that the framework captures disease-associated biological age deviation in a heterogeneous manner across organs, with the strongest PDAC-associated perturbation localized to the pancreas.

Importantly, these findings should be interpreted as diagnostic external clinical validation, not as external validation of pre-diagnostic risk. The PANORAMA analysis demonstrates that the model is sensitive to disease-associated variation in an external clinical cohort and that this sensitivity is biologically heterogeneous across organs; however, it does not replace the primary in-house pre-diagnostic analyses that underlie the main conclusions of the study. This framing is also consistent with the original PANORAMA analysis plan, which positioned the cohort as a supplementary external clinical stress test rather than as the primary validation set for pre-diagnostic disease association.

**Supplementary Note 2. Chronological age modifies BAG-associated disease risk**

Biological age gap showed marked age-dependent effect modification in its association with future disease risk. For the ensemble biomarker, the relative hazard associated with a higher age gap was strongest in younger individuals, peaked around midlife, and then declined steadily with advancing chronological age, reaching null in the early 70s (**Figure S2a**). The significant interaction term (**p_interaction < 0.001**) supports the conclusion that chronological age attenuates the association between ensemble biological age gap and disease risk. Importantly, the attenuation occurred within the age range containing the largest concentration of subjects and events, rather than being restricted to sparsely sampled extremes.

A similar pattern was observed across organ-specific biological age gaps (**Figure S2b**). At age 45, most biomarkers showed elevated hazard ratios, indicating that higher biological age relative to chronological age was associated with increased future disease risk. By age 75, these associations had generally weakened toward the null. The extent of attenuation differed by organ: liver and kidney attenuated earliest, ensemble and aorta showed intermediate persistence, and pancreas retained the strongest association across age, with the fitted HR remaining above 1 throughout the modeled range. Although spleen exhibited a statistically significant interaction, its overall effect size appeared weaker and closer to the null than the other biomarkers. These findings suggest that biological age gap captures a disease-relevant aging signal that is most pronounced earlier in the aging trajectory and becomes less discriminative with increasing chronological age.

We observed a significant interaction between chronological age and both ensemble and organ-specific BAG, such that the hazard ratio associated with a given BAG increment declined with advancing chronological age. This pattern is consistent with emerging evidence across several biological-aging biomarker domains, although direct studies of chronological-age modification of BAG for incident disease endpoints remain limited. For example, Kuo et al. reported that associations of some baseline epigenetic clocks, specifically Dunedinm38 and DunedinPACE, with mortality were weaker in older than in younger adults, and Dong et al. reported that chronological age significantly modified the association between vascular age residuals and cardiovascular events, with stronger predictive value in middle-aged than in older adults [[3,37]].

One possible explanation is that, with increasing chronological age, higher background event rates, survival selection, multimorbidity, and competing mortality reduce the relative discriminative value of a fixed BAG increment. Consistent with this broader age-attenuation pattern, the relative mortality effect of frailty weakens with advancing age, the mortality burden associated with a given multimorbidity count is markedly larger in younger than in older adults, and pulse-pressure-related relative risk declines with age even as absolute risk rises [[38,39,40]]. A similar age-related attenuation has also been reported for self-rated health as a mortality predictor in some cohorts, although findings are not fully uniform across populations [[41]].

Importantly, attenuation of relative effect does not imply lack of clinical utility in older adults. Biological-age measures can still carry prognostic information beyond chronological age, but our findings suggest that their relative risk-stratification value may be greatest earlier in the aging trajectory [[7]]. More broadly, recent biomarker-of-aging frameworks distinguish between measures of accumulated biological aging and measures of pace of aging, implying that age-aware BAG modeling, chronological-age interaction terms, age-stratified calibration, and complementary pace-based metrics may improve clinical translation [[5]]. Together, these findings suggest that organ-specific BAG may be especially informative for prevention-oriented risk stratification in younger and midlife adults, while remaining interpretable in older adults through an absolute-risk framework.

### Supplementary Table

| Criteria | Cohort#1 (Control) | Cohort #2 (Disease) | Cohort#3 (PANORAMA) | Cohort#4 (External Control) |
|---|---|---|---|---|
| 40 and less | 352(1.6%) | 0(0%) | 109(5.6%) | 1(0.8%) |
| 40-60 | 8743(40.2%) | 9808(21.9%) | 451(23.1%) | 30(25%) |
| 60-80 | 10061(46.2%) | 24750(55.2%) | 1177(60.3%) | 74(61.7%) |
| 80 and more | 2603(12.0%) | 10287(22.9%) | 222(11.4%) | 15(12.5%) |
| No. of records | 21759 | 44844 | 1959 | 120 |
| Male | 11467(52.7%) | 19782(44.1%) | 1060(54.3%) | 63(52.5%) |
| Female | 10292(47.3%) | 25062(55.9%) | 899(45.7%) | 57(47.5%) |
| No. of Subjects | 11917 | 18900 | 1945 | 120 |
| Male | 6191(52.0%) | 8612(45.6%) | 1052(54.1%) | 63(52.5%) |
| Female | 5726(48.0%) | 10288(54.4%) | 893(45.9%) | 57(47.5%) |

**Table S1. Demographic Information of Included Data Cohorts**

| Model | MAE | 95% CI | R2 | PCC | Age Range |
|---|---|---|---|---|---|
| **Cross Validation** | | | | | |
| **Ensemble** | **3.68** | **3.62-3.75** | **0.90** | **0.95** | **20-100** |
| **Aorta** | 5.85 | 5.74-5.96 | 0.74 | 0.87 | 40-100 |
| **Pancreas** | 5.94 | 5.83-6.05 | 0.73 | 0.87 | 40-100 |
| **Liver** | 5.56 | 5.46-5.67 | 0.76 | 0.88 | 40-100 |
| **Kidney** | 5.64 | 5.54-5.75 | 0.76 | 0.89 | 40-100 |
| **Spleen** | 5.37 | 5.27-5.47 | 0.78 | 0.89 | 40-100 |
| **Internal Validation** | | | | | |
| **Ensemble** | **4.06** | **3.98-4.14** | **0.84** | **0.92** | **40-100** |
| **Kerber et al. [3] (Inhouse trained)** | 4.90 | 4.52-5.29 | 0.79 | 0.89 | 40-100 |
| **External validation** | | | | | |
| **Ensemble(Multi-Center)** | **4.58** | **4.07-5.10** | **0.78** | **0.88** | **30-90** |
| **Literatures** | | | | | |
| **Kerber et al. [3]** | 5.76 | 5.07-6.45 | 0.71 | - | 40-80 |
| **Wang et al. [11]** | 5.67 | - | 0.81 | 0.90 | 20-100 |
| **Lin et al. [12]** | 5.15 | 4.81-5.48 | 0.74 | - | - |

**Table S2. Result of Age Prediction model on alignment with Controlled population**

### Supplementary Figure

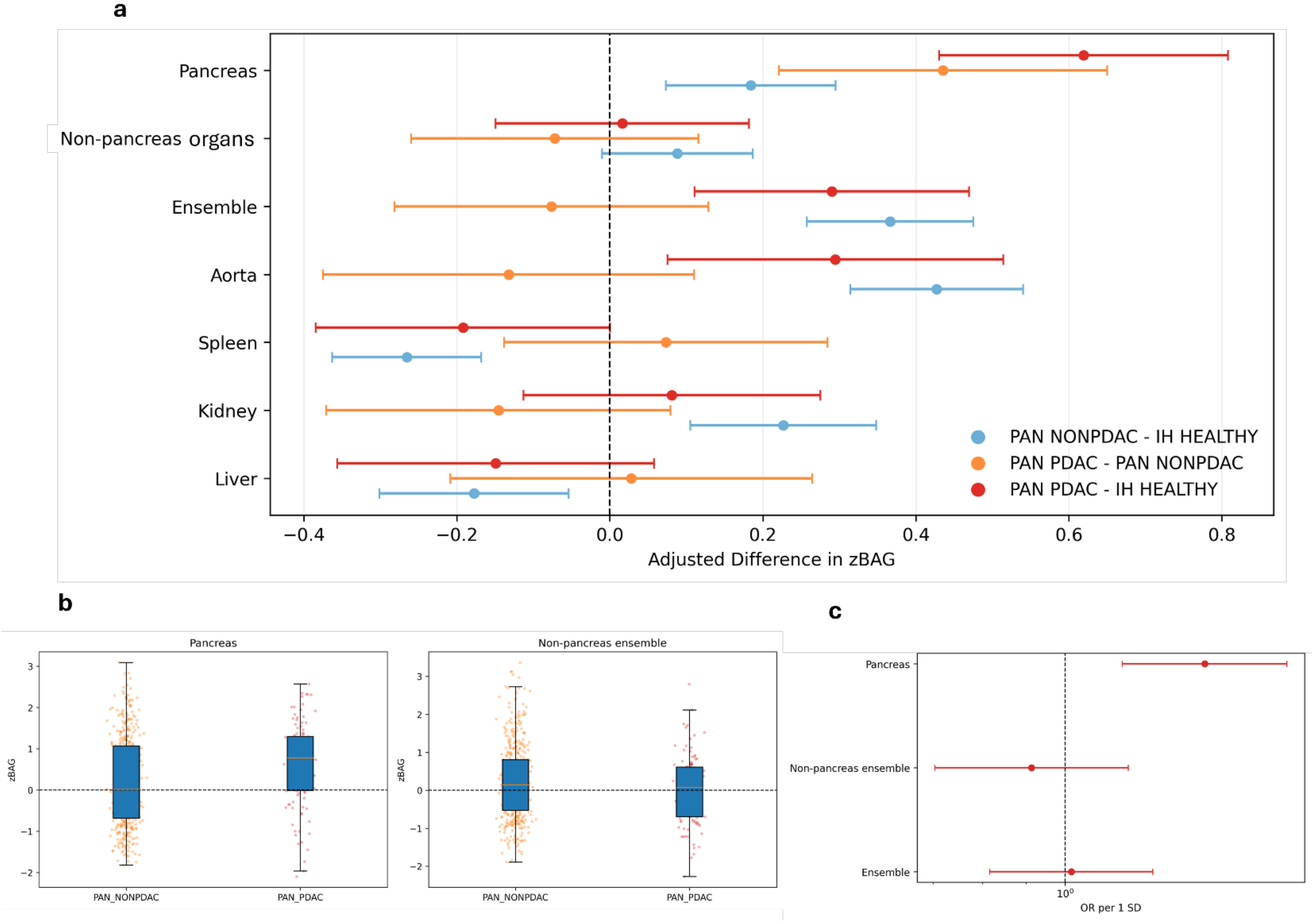


**Figure S1 | External validation of biological age deviation in the Pancreatic Ductal Adenocarcinoma (PDAC) cohort in PANORAMA.**

a) adjusted differences in BAG (zBAG) with 95% confidence intervals for the planned contrasts PAN_NONPDAC − IH_HEALTHY, PAN_PDAC − PAN_NONPDAC and PAN_PDAC − IH_HEALTHY across the pancreas, non-pancreatic organs, ensemble estimate and individual organs. The dashed vertical line indicates zero difference. Bottom left and middle, distributions of pancreatic and non-pancreatic ensemble zBAG in PAN_NONPDAC and PAN_PDAC subjects. c) Odds ratios for PDAC per 1 s.d. increase in zBAG from PANORAMA-only logistic models. PAN_NONPDAC subjects already differed from the in-house healthy reference, whereas PDAC cases showed an additional increase in pancreatic zBAG relative to PAN_NONPDAC controls, with weaker non-pancreatic and ensemble signals.

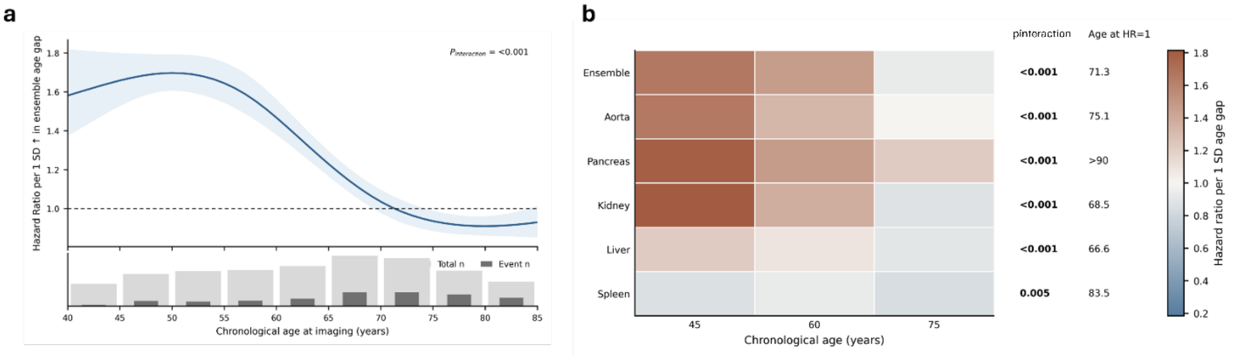


**Figure S2 | Chronological age attenuates the association between biological age gap and future disease risk.**

a) Age-dependent association of ensemble biological age gap with disease risk. The blue line shows the model-estimated hazard ratio (HR) per 1 SD increase in ensemble age gap across chronological age, with shaded 95% confidence intervals. The dashed line indicates the null (HR = 1). $P_{interaction}$ denotes the significance of age-dependent effect modification. The lower strip shows total sample counts (light gray) and event counts (dark gray) across age bins. b) Organ-level summary of age-dependent attenuation. Heatmap cells show the predicted HR per 1 SD increase in biological age gap at ages 45, 60, and 75 years for the ensemble and organ-specific models. Warmer colors indicate stronger associations, whereas lighter colors indicate attenuation toward the null. Pinteraction denotes the significance of the age-by-BAG interaction term, and Age at HR = 1 denotes the modeled age at which the association reaches the null. Associations were generally stronger at younger ages and weaker at older ages, with pancreas showing the most persistent effect.

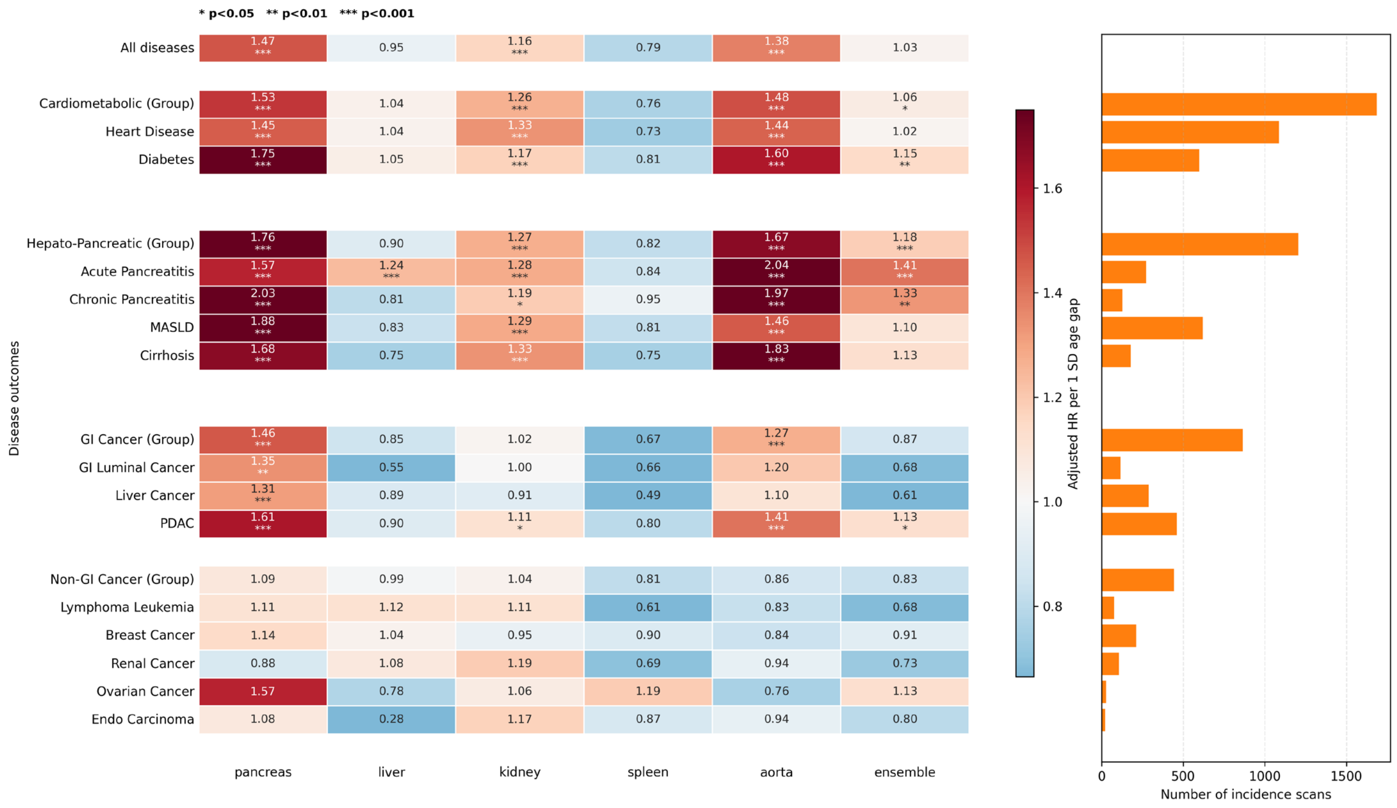


**Supplementary Figure S3 | Sensitivity analysis of organ-specific biological age gap associations with future disease risk in adults aged 20–80 years.**

Heatmap shows adjusted hazard ratios (HRs) per 1-s.d. increase in organ-specific or ensemble biological age gap (BAG) from Cox proportional hazards models across pooled and disease-specific outcomes in the expanded 20–80-year analysis cohort. Models were adjusted for chronological age, sex, and body mass index. Columns correspond to pancreas, liver, kidney, spleen, aorta, and ensemble BAG. Warmer colors indicate higher HRs, and cooler colors indicate lower HRs. Asterisks denote significance (*p < 0.05, **p < 0.01, ***p < 0.001). The bar plot on the right shows the number of pre-diagnostic incidence scans contributing to each outcome. Compared with the focused 20–60-year analysis, pancreas and aorta BAG retained broad disease associations, particularly for cardiometabolic and hepato-pancreatic outcomes, whereas liver, spleen, and ensemble signals were attenuated, consistent with chronological-age-dependent weakening of relative BAG-associated risk.
